# Dysfunctional cortical gradient topography in treatment resistant major depression

**DOI:** 10.1101/2022.06.16.22276402

**Authors:** Lorenzo Pasquini, Susanna L. Fryer, Stuart J. Eisendrath, Zindel V. Segal, Alex J. Lee, Jesse A. Brown, Manish Saggar, Daniel H. Mathalon

## Abstract

**Background:** Treatment-Resistant Depression (TRD) refers to patients with major depressive disorder who do not remit after two or more antidepressant trials. TRD is common and highly debilitating, but its neurobiological basis remains poorly understood. Recent neuroimaging studies have revealed cortical connectivity gradients that dissociate primary sensorimotor areas from higher-order associative cortices. This fundamental topography determines cortical information flow and is affected by psychiatric disorders. We examined how TRD impacts this hierarchical cortical organization.

**Methods:** We analyzed resting-state fMRI data from a mindfulness-based intervention study in 56 TRD patients and 28 healthy controls. Using novel gradient extraction tools, measures of cortical gradient dispersion within and between functional brain networks were derived, compared across groups, and associated with graph theoretical measures of network topology. Within TRD, baseline cortical gradient dispersion measures were correlated with baseline clinical measures (anxiety, depression, mindfulness), as well as with changes in these measures following treatment with either mindfulness-based therapy or a health enhancement program.

**Results:** Cortical gradient dispersion was reduced within major intrinsic brain networks in TRD. Reduced cortical gradient dispersion correlated with increased network modularity assessed through graph theory-based measures of network topology. Lower dispersion among Default Mode Network regions, a transmodal system linked to depression symptomatology, related to current levels of trait anxiety, depression, and mindfulness, but not to changes in these domains following treatment.

**Conclusions:** Our findings reveal widespread alterations in cortical gradient architecture in TRD, implicating a significant role for the Default Mode Network in mediating depression, anxiety, and lower mindfulness in patients.

## Introduction

Major depression is a common, debilitating disorder and among the leading causes of disability worldwide (1). Although several treatment options are available for depression, a significant number of patients do not improve despite adequate antidepressant trials (2). Patients that, after repeated treatments, fail to reach acceptable levels of functioning and well-being, eventually present with treatment-resistant depression (TRD), a condition associated with a significant social and economic burden (2,3). TRD is often defined as the failure to remit after at least two antidepressant trials of adequate dose and duration (2,3). A consensus characterization of TRD, however, has yet to be achieved, partly due to poor understanding of its neurobiological basis and a lack of reliable diagnostic biomarkers (4,5).

Resting-state fMRI (rs-fMRI) is a neuroimaging modality commonly used to measure functional connectivity of brain networks in terms of correlated spontaneous activity among distant brain regions (6,7). This method has proven useful in revealing altered functional connectivity within and between various limbic and higher-order brain networks in depression (5,8,9). Crucially, several depression studies have linked maladaptive self-referential processes, such as rumination and emotional dysregulation, to functional alterations of the Default Mode Network (DMN) (9–11), a transmodal system comprising the medial prefrontal, medial temporal, medial parietal, and angular cortices (12,13).

Fundamental principles in behavioral neurology and recent neuroimaging studies provide convergent support for a hierarchical cortical organization that separates primary sensorimotor systems from transmodal associative areas (14–16). Cortical microstructure, connectivity, and gene expression findings point to dominant sensorimotor-to-transmodal gradients organizing the propagation of sensory inputs from primary areas into transmodal regions along multiple cortical relays (14,15,17). This large-scale brain system organization anchors the DMN at one end of the hierarchy with respect to primary sensorimotor areas, capturing a functional topography that enables the transition from perception to more abstract cognitive functions (9–11). Several neuropsychiatric disorders, including major depression (18), cognitive vulnerability to depression (19), and autism (17), profoundly impact connectivity-based cortical gradient organization. Remarkably, major depression both disrupts global topography and produces focal alterations of cortical gradients among primary sensory and transmodal regions involved in high-order cognitive processing (18).

Accordingly, we hypothesized that TRD would impact hierarchical brain network organization and that functional deficits affecting the DMN would predict current and future symptoms of depression following group treatment with either mindfulness-based cognitive therapy (MBCT) or a health enhancement program (HEP). We applied a novel gradient decomposition technique (20) to baseline rs-fMRI data from 56 TRD patients subsequently randomized to MBCT or HEP, and from 28 healthy controls (HC). This approach was leveraged to test the hypothesis that TRD, relative to HC, involves perturbation of hierarchical gradients among “canonical” large-scale brain networks (21), and to further contextualize the results using graph theoretical metrics of network topology, specifically nodal degree (22).

## Materials and Methods

### Subjects

All participants or their surrogates provided written informed consent prior to participation in accordance with the declaration of Helsinki. The University of California San Francisco Committee on Human Research approved the study.

An initial cohort of 59 TRD patients were enrolled in randomized controlled behavioral intervention study that included baseline and post-treatment fMRI sessions, and 30 HC were recruited to provide normative baseline fMRI data. Participants were recruited from outpatient psychiatry and general medicine clinics at the University of California San Francisco (UCSF), the outpatient psychiatry clinic at Kaiser Permanente in San Francisco, and through fliers, Craigslist advertisements, and clinical referrals, as described previously (23,24). TRD patient eligibility screening was completed in person. Eligible patients met Structured Clinical Interview for DSM-IV-TR Axis I (SCID-I/P) (25) criteria for major depression and had a Hamilton Depression Severity Rating Scale (HAMD-17) score of 14 or greater. Furthermore, to qualify as TRD, patients had to be taking antidepressant medication with evidence of two or more adequate trials prescribed during the current episode as assessed with the Antidepressant Treatment History Form (26). Patients were excluded for the following: lifetime history of bipolar disorder, schizophrenia, or any psychotic disorder; substance abuse or dependence within three months of study onset; currently suicidal, dangerous to others, or self-injurious; undergoing psychotherapy that they were unwilling to discontinue during the eight-week treatment portion of the study (further details below); or a score of <25 on the Mini Mental Status Exam (27).

The HC group was matched to the TRD group on age, gender, and handedness and had no history of a major Axis I psychiatric disorder, neurological illness, or current use of psychotropic medication. Participants were required to be at least 18 years old, fluent in English, have no MRI contraindications, and to have normal or corrected-to-normal vision.

For each participant, we additionally assessed depressive symptoms through the Quick Inventory of Depression Symptomatology (QIDS-SR16) (28) and the Nolen-Hoeksema’s Response Styles Questionnaire (RSQ22) (29); levels of mindfulness were assessed with the Five Facet Mindfulness Questionnaire (FFMQ) (30); and levels of state and trait anxiety were assessed through the State-Trait Anxiety Inventory (STAI trait and state) (31). Study participants self-reported race and ethnicity. Sex, handedness, and years of education were also reported.

From the initially recruited sample, two HCs and three TRD patients had to be excluded based on excessive head movement in the scanner (see details below), resulting in the final analyzed sample of 56 TRD and 28 HC participants (Table 1).

**Table 1.** Participants’ demographic and clinical characteristics at baseline. Mean and standard deviation in brackets. ^&^Chi-square test. FD = framewise head displacement; FFMQ = Five Facet Mindfulness Questionnaire; HAMD-17 = Hamilton Depression Rating Scale; HC = healthy control; MAOI = monoamine oxidase inhibitors; MDE = major depressive episode; QIDS-SR16 = Quick Inventory of Depression Symptomatology; RSQ22 = Nolen-Hoeksema’s Response Styles Questionnaire; SNRI = selective and norepinephrine reuptake inhibitors; SRI = selective reuptake inhibitors; SSRI = selective serotonin reuptake inhibitors; STAI = State-Trait Anxiety Inventory; TCA = tricyclic antidepressants; TRD = treatment resistant major depression.

|  | **HC**<br>**(n=28)** | **TRD**<br>**(n=56)** | **T** | **p** |
| --- | --- | --- | --- | --- |
| Age in years | 45.4 (9.3) | 42.9 (9.9) | 1.14 | 0.260 |
| Female | 20 | 44 | 0.21<sup>&</sup> | 0.651 |
| Handedness ambidextrous/left/right | 1/2/25 | 2/5/49 | 0.08<sup>&</sup> | 0.962 |
| Education in years | 16.9 (2.5) | 16.1 (2.1) | 1.57 | 0.123 |
| Hispanic-Latino | 4 | 4 | 0.40<sup>&</sup> | 0.529 |
| Asian/Black/Other/White | 1/2/0/25 | 6/4/1/45 | 12.38<sup>&</sup> | <0.01 |
| Mean FD in mm | 0.23 (0.10) | 0.25 (0.11) | -1.01 | 0.316 |
| Age of MDE onset in years | - | 20.8 (10.1) | - | - |
| Number of MDEs | - | 3.6 (2.5) | - | - |
| Current onset duration in months | - | 85.6 (110.5) | - | - |
| Number of trials | - | 2.9 (1.3) | - | - |
| Concurrent medication at baseline |  |  |  |  |
| Antidepressants | - | 56 (100.0%) | - | - |
| Mood stabilizers | - | 8 (14.3%) | - | - |
| Sedatives | - | 19 (33.9%) |  |  |
| Stimulants | - | 13 (23.2%) | - | - |
| Antipsychotics | - | 1 (20.0%) | - | - |
| Other | - | 1 (1.8%) | - | - |
| Clinical questionnaires |  |  |  |  |
| HAMD-17 | 1.6 (1.3) | 17.4 (2.7) | -35.5 | <0.001 |
| QIDS-SR16 | 2.6 (1.4) | 14.9 (3.7) | -21.6 | <0.001 |
| STAI trait | 27.6 (5.8) | 60.1 (8.5) | -19.6 | <0.001 |
| STAI state | 26.5 (7.8) | 56.3 (9.8) | -14.5 | <0.001 |
| RSQ22 | 31.8 (9.0) | 59.7 (11.0) | -12.0 | <0.001 |
| FFMQ | 157.2 | 106.1 | 12.0 | <0.001 |

### Protocol

TRD patients were part of a randomized controlled trial comparing MBCT to a HEP as adjunctive treatments to ongoing antidepressant medication. From our final sample of 56 TRD patients, 27 underwent MBCT and 28 underwent HEP. Details regarding treatment programs and randomization procedures were described previously (23,24). Briefly, MBCT involved guided meditations and exercises intended to help participants identify cognitive distortions, disengage from rumination, and use nonjudgmental present-moment awareness (26). HEP involved physical exercise, functional movement, music therapy, diet education, and guided imagery intended to promote health and improve mood (33). Both treatment groups met for eight weeks in groups of 6–12 once a week for 135 min. Patients were assessed with rs-fMRI at baseline and following intervention, while HC were assessed at baseline and did not undergo treatment. Only baseline rs-fMRI data from TRD and HC participants are analyzed in the present study. TRD patients underwent clinical assessments at baseline and at weeks 8, 24, 36, and 52 (23,24).

### Neuroimaging data acquisition and preprocessing

Neuroimaging data were acquired on a Siemens 3-T TIM TRIO scanner located at the UCSF Neuroimaging Center. A high-resolution anatomical scan was acquired using a 3-D MP-RAGE sequence, with scan time 5 min 17 s, flip angle 9 degrees, FOV = 220 mm, 160 slices per slab, 1.2 mm thick, no gap, TR = 2.30 s, TE = 2.94 ms. Functional scans were acquired using an EPI-BOLD sequence, TR = 2, TE= 30 ms, FoV = 220 MM, flip angle = 77 degrees, bandwidth = 2298 Hx/pixel, matrix = 64 x 64. 30 slices (3 mm thick, 1-mm gap). Scans were acquired in an axial-oblique plane, parallel to the anterior-posterior commissure (AC-PC) line. Participants were instructed to rest with eyes open during the 5 min and 24 s EPI-BOLD functional sequence.

The software fMRIPrep (https://fmriprep.org/en/stable/) (33) was used for subsequent neuroimaging data preprocessing. Anatomical MP-RAGE images were corrected for intensity non-uniformity, skull-stripped, and segmented for cerebrospinal fluid, white matter, and gray matter. Volume-based spatial normalization to MNI standard space was performed through nonlinear registration of the MP-RAGE with the T1-weighted MNI template brain (CBM152). The first five functional image volumes were removed to allow for scanner equilibration. A mean reference volume and its skull-stripped version were generated, then co-registered to the structural reference using affine registration. Head-motion parameters (transformation matrices and the six corresponding rotation and translation parameters) were estimated and used to compute framewise head displacement time series. Functional images were slice-time corrected, realigned, and normalized to MNI standard space applying the structural transformation matrix to the co-registered functional data. The resulting volumes with 2 mm^3^ isotropic resolution were spatially smoothed with a 6 mm radius Gaussian kernel and bandpass filtered in the 0.008–0.15 Hz frequency range. Nuisance parameters in the preprocessed data were estimated for the cerebrospinal fluid and white matter. Additional nuisance parameters included the three translational and three rotational motion parameters, the temporal derivatives of the previous eight terms (white matter/cerebrospinal fluid/six motion time series), and the squares of the previous 16 terms (34,35). Nuisance parameters were filtered for the same frequency range as rs-fMRI data and regressed out from the filtered rs-fMRI data (34,35). The denoised data were used in subsequent analyses. Subjects were included only if their mean framewise head displacement in the scanner (34,35) was below the threshold of 0.55 mm recommended in previous work (36). Global signal regressed rs-fMRI data were also generated using the time series extracted from a whole-brain mask and used for control analyses.

### Functional connectivity gradients

The Schaefer Atlas (37) was used to derive rs-fMRI activity time series for 400 cortical regions (Figure 1A). This data-driven atlas, derived from 1498 healthy individuals, exploits local gradients in functional connectivity, while maximizing the similarity of rs-fMRI time courses within a parcel. The resulting cerebral cortex parcellations are functionally and connectionally homogeneous and display a one-to-one correspondence to major intrinsic brain networks (21) (Figure 1B). Pearson’s correlation was applied to the regional activity time series to derive individual functional connectivity matrices (Figure 1Ca) and group-mean functional connectivity matrices for HC and TRD participants (Figure S1).

**Figure 1.**
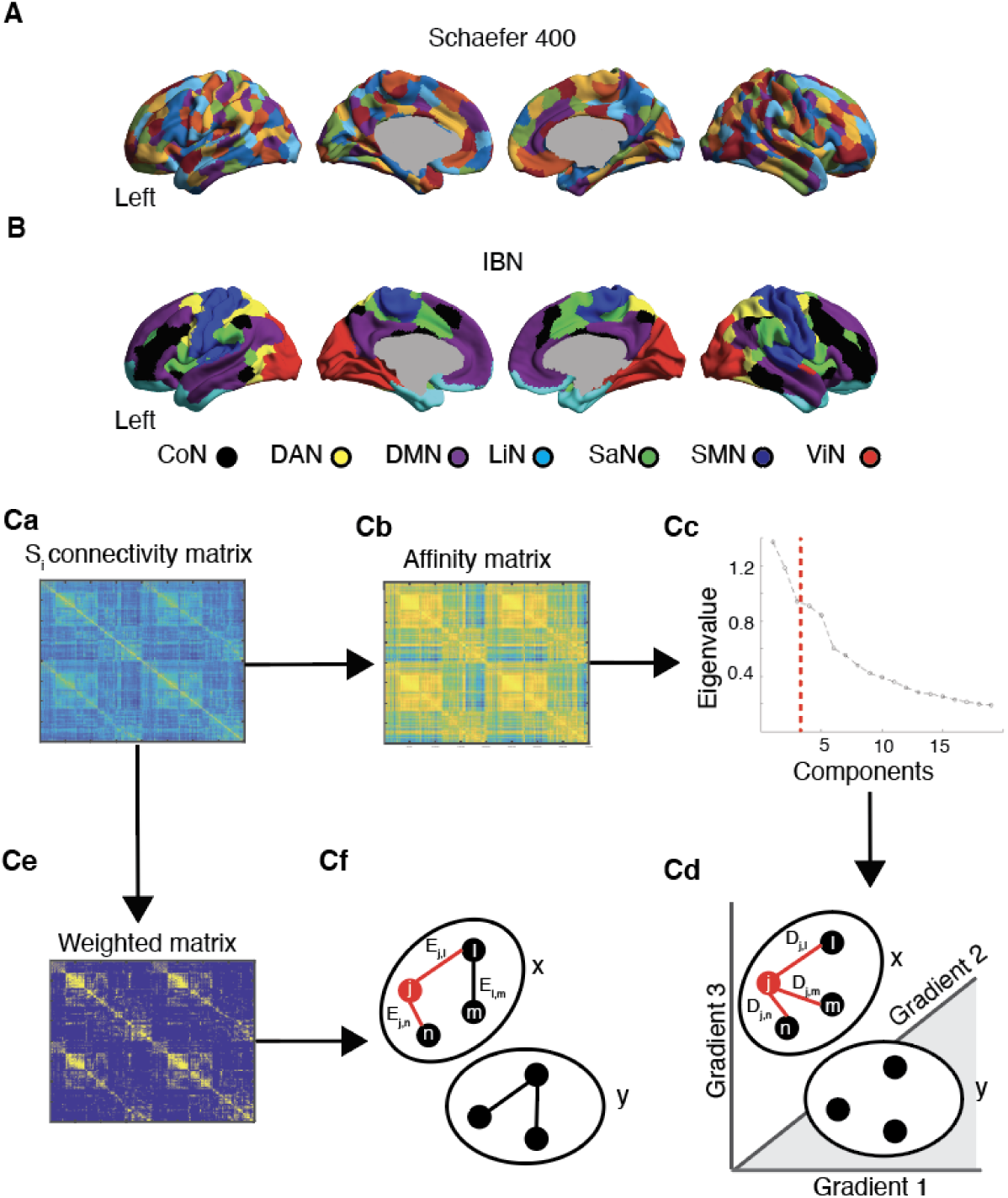
Analytic pipeline. **(A)** 400 nodes from the Schaefer Atlas, each overlapping with a specific intrinsic brain network (IBN) **(B)**, were used to derive functional connectivity matrices using rs-fMRI data of HCs and patients with TRD. **(Ca).** Individual connectivity matrices (Si) went through two distinct processing pipelines. To derive cortical connectivity gradients (upper stream), individual connectivity matrices were transformed to affinity matrices using cosine similarity (**Cb**) and Laplacian decomposition was used to derive three primary connectivity gradients, which explained most of the variance in the data (**Cc**). The position of an individual node belonging to a specific intrinsic brain network (e.g. Network x) was used to derive a topographical measure of nodal dispersion (**Cd**), reflecting the average Euclidean distance in gradient space between a node and all other nodes belonging to the same network. Individual connectivity matrices were also leveraged to derive topological measures of nodal degree (lower stream). Connectivity matrices were weighted by binarizing at a connectivity threshold of 0.35 (**Ce**). For each node within a network, we assessed the level of degree by counting the edges of this node to all other nodes within a network and dividing by the total amount of edges (**Cf**). CoN = Control Network; DAN = Dorsal Attention Network; DMN = Default Mode Network; HC = healthy controls; LiN = Limbic Network; SaN = Salience Network; SMN = Sensorimotor network; TRD = patients with treatment resistant depression; ViN = Visual Network.

The diffusion embedding approach (14,15), as implemented by the toolbox BrainSpace (https://brainspace.readthedocs.io/en/latest/pages/getting_started.html) (20), was then applied to the HC group mean functional connectivity matrix to estimate connectivity gradients. Briefly, the top 10% strongest functional connections were retained for each parcel, referred to hereafter as a node, and cosine similarity was calculated between each pair of nodes to generate a dissimilarity matrix (Figure 1Cb) (38,39). Diffusion map embedding, a data reduction technique, was then applied to decompose the functional connectome into primary components, referred to as gradients, explaining most of the variance in connectivity (Figure 1Cc). These gradients discriminate across levels of the cortical hierarchy (i.e., sensory processing versus higher-order cognition), whereas node-specific gradient values reflect the similarity in connectivity along this sensory-transmodal axis. An identical approach was used to derive connectivity gradients from the TRD group mean connectivity matrix and from the connectivity matrices of individual participants. The resulting gradient maps were subsequently aligned to the gradients derived at the group-level in HCs using iterative Procrustes rotation, therefore enabling comparisons across individual embedding solutions (17,21,40).

### Nodal dispersion

For each participant, we then derived a measure of within-network nodal dispersion. We plotted the first three connectivity gradients – since these explained most of the underlying variance (see also elbow plot in Figure 1 Cc) – against each other to derive a three-dimensional manifold in which we calculated the Euclidean distance between nodes belonging to the same intrinsic brain network (42) (Figure 1Cd). Nodal dispersion was derived for each node belonging to a specific intrinsic brain network and averaged across nodes within intrinsic brain networks, yielding a final estimate of within-network nodal dispersion for each participant. We performed several control analyses to assess the impact of methodological parameters on our analyses (see Supplement). Further, we derived a measure of between-network nodal dispersion calculated as the Euclidean distance between network centroids (i.e., the arithmetic mean in gradient space of all nodes belonging to the same network).

### Nodal degree

In parallel to the novel connectivity gradient approach, we also derived a traditional measure of within-network nodal degree for all participants (22) by using the publicly available Brain Connectivity Toolbox (https://sites.google.com/site/bctnet/).

Nodal degree is a widely used measure of network topology commonly derived using graph-theoretical approaches (22). Briefly, individual connectivity matrices were thresholded for correlation values below 0.35 and binarized (Figure 1Ce). For control analyses, measures of nodal degree were derived also for connectivity thresholds of 0.45 and 0.25. Weighted connectivity matrices were used to count the number of surviving edges between a specific node within a network and all other nodes within the same network (Figure 1Cf). The sum of surviving edges for a node was then divided by the total amount of edges within the network. Nodal degree measures were derived for each single node in a network and averaged across nodes in the same network. This procedure resulted in a measure of within-network nodal degree reflecting the level of integration between nodes belonging to the same network.

### Statistical analyses

In house MATLAB R2021a (https://www.mathworks.com/products/matlab.html) and R 4.1.1 (https://www.r-project.org/) scripts were used to perform the statistical analyses. For more details, see Supplementary Methods.

## Results

### Cortical connectivity gradients in HCs and TRD patients

We applied a diffusion gradient approach separately on rs-fMRI-based connectivity data from HCs and TRD patients to derive cortical connectivity gradients reflecting processing hierarchies spanning sensory and transmodal areas (Figure 2 and Figure S2A). We next describe the first three principal gradients derived from rs-fMRI data of HCs, since these explained most of the variance in functional connectivity (elbow plot in Figure 1 Cc). Gradient 1 anchored sensorimotor areas at its positive extreme, while regions belonging to the DMN were located at the opposite, negative extreme (Figure 2A-B). The DMN occupied the negative extreme on Gradient 2, while visual-sensory areas populated the positive end of this gradient (Figure 2A-B). Notably, these first two connectivity gradients overlap with previously reported gradients in functional connectivity, structural connectivity, myelin density, and genetic expression (14,15), which consistently separate sensory processing regions from transmodal areas of the DMN. Gradient 3 showed a more complex pattern, segregating regions of the Dorsal Attention Network from regions belonging to the Salience Network, potentially reflecting a higher-order, attention-related gradient separating regions attending to externally presented cues (43) from regions devoted to processing visceral and interoceptive information (44,45). Similar fundamental properties of hierarchical brain organization were found in patients with TRD after aligning the principal connectivity gradients of patients to those of HCs (Figure 2C-D), in support of the notion that cortical gradients reflect fundamental properties of brain topography in both health and disease (14,15,17,18,42). Of note, the elbow plot in Figure 1Cc suggested that also Gradients 4-6 contributed to a significant amount of explained variance, although to a lesser degree when compared to the first three gradients. These cortical gradients displayed less discernable patterns of regional variation, separating sensory areas from regions belonging to cingulo-opercular and frontoparietal networks (Figure S2).

**Figure 2.**
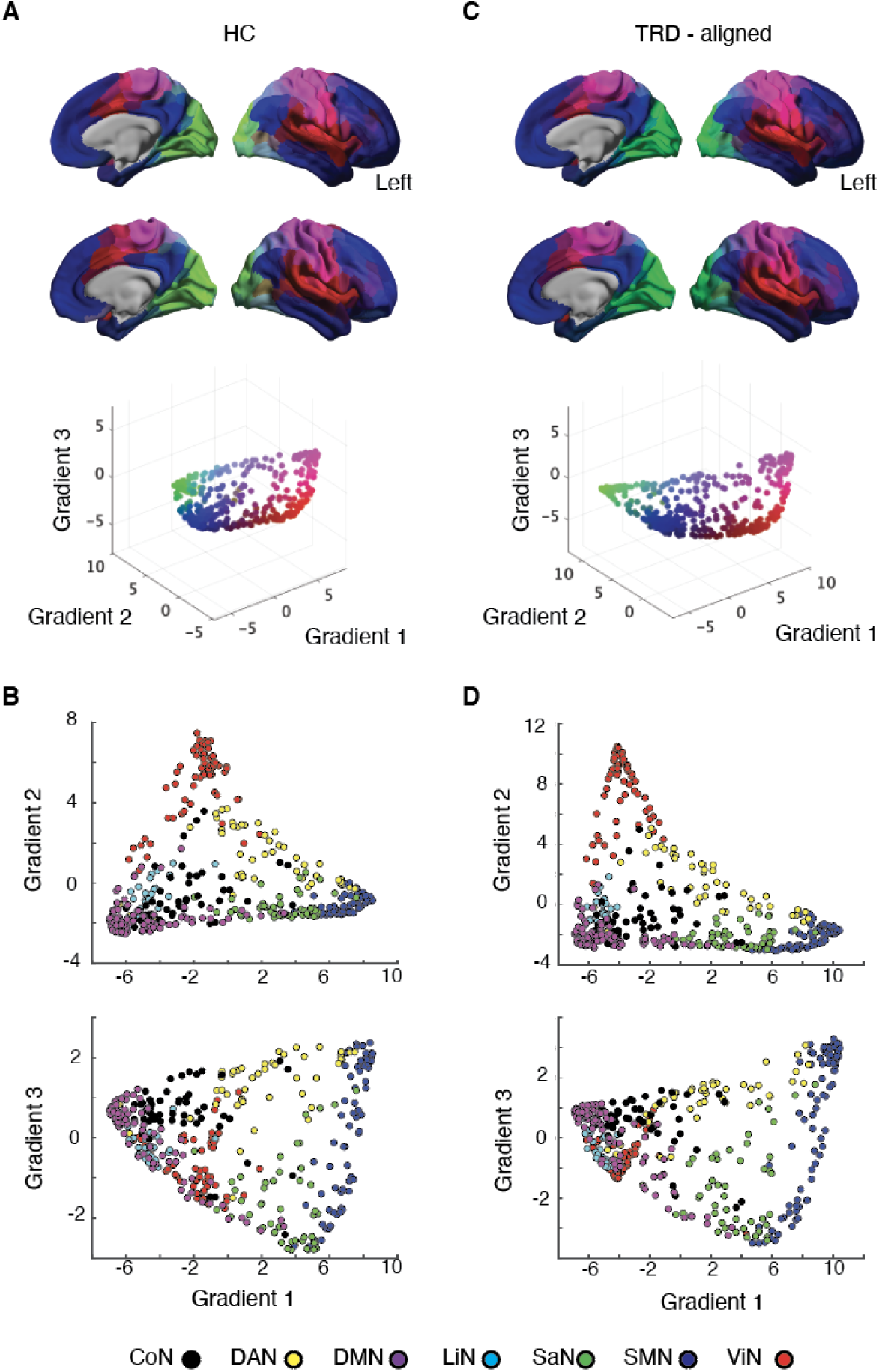
Cortical connectivity gradients. **(A)** Cortical connectivity gradients of HCs projected into cortical surface. The three-dimensional scatterplot below shows how individual nodes distribute along the first three gradients. Colors reflect the loadings of nodes on individual gradients. For example, the sensorimotor cortex appears purple and regions overlapping with the DMN appear blue, reflecting that these systems respectively anchor the extremes of Gradient 1. **(B)** Scatterplots reflecting how nodes belonging to distinct intrinsic brain networks align along cortical gradients in HC. **(C)** Cortical connectivity gradients of patients with TRD aligned to the gradients of HCs following Procrustes rotation. **(D)** Scatterplots reflecting how nodes belonging to distinct intrinsic brain networks align along cortical gradients in patients with TRD. CoN = Control Network; DAN = Dorsal Attention Network; DMN = Default Mode Network; HC = healthy controls; LiN = Limbic Network; SaN = Salience Network; SMN = Sensorimotor network; TRD = patients with treatment resistant depression; ViN = Visual Network.

### Within-network nodal dispersion

Node-level gradient comparisons (p<0.05, uncorrected) revealed increased gradient scores in TRD patients in sensory and early transmodal regions, such as the ventromedial occipital and posterior inferior temporal cortices, together with decreased gradient scores in transmodal areas including the precuneus, the medial prefrontal, and cingulate cortices (Figure 3A). We then derived a measure of within-network nodal dispersion (Figure 1Cd), reflecting how closely nodes belonging to the same intrinsic brain network are located in the topographical three-dimensional gradient space (42). All networks, except for the Salience and Sensorimotor Networks, showed reduced within-network nodal dispersion in TRD patients compared to HCs (Figure 3B; p<0.05, FDR corrected for multiple comparisons). We explored the relationship between within-network nodal dispersion in TRD patients and confounds such as mean frame-wise head displacement during scanning and demographic variables, including age and sex, using multiple linear regression models (Table S1; p<0.05, uncorrected), which revealed no significant associations. Of note, adding Gradients 4-6 when computing measures of within-network nodal dispersion affected group differences, with only the Visual and Control Networks showing significantly reduced within-network nodal dispersion in TRD (Table S2; p<0.05, FDR corrected for multiple comparisons). We performed additional control analyses to assess the impact of methodological parameters on group differences in within-network nodal dispersion. We found similar cortical gradients as those reported in the main findings regardless of whether we used: (1) global signal regression; (2) higher or lower parcellated atlases; (3) gradient decomposition through Laplacian embedding; (4) or angular normalization to generate the dissimilarity matrices (Figure S2 C-F). While data preprocessed with global signal regression or higher atlas parcellation consistently revealed decreased within-network nodal dispersion in TRD, group differences were affected when using a lower parcellated atlas, Laplacian embedding, or angular normalization (Table S2). Finally, we analyzed whether TRD also affected cortical hierarchies between networks in addition to within-network gradient organization. We derived a measure of between-network nodal dispersion that revealed reduced nodal dispersion in TRD between the Sensorimotor and the DMN, between the Salience and the DMN, and between the Control and Dorsal Attention Network, although none of these findings survived FRD correction for multiple comparisons (Figure S3; p<0.05, uncorrected).

**Figure 3.**
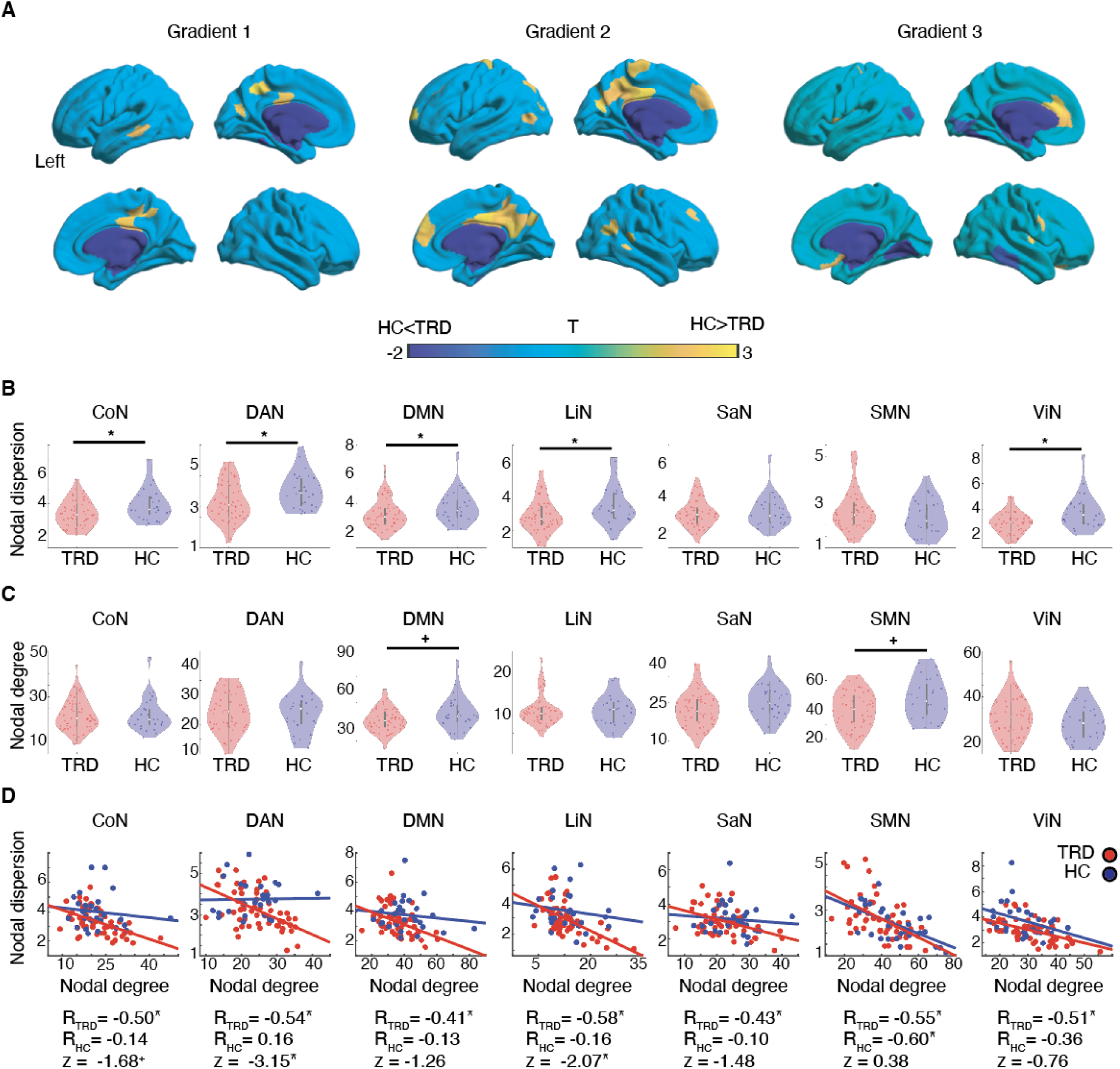
Nodal dispersion and nodal degree. **(A)** Node-wise statistical comparisons between HCs and TRD, with increases/decreases in TRD shown in cold/warm colors (p<0.05 uncorrected). **(B)** Violinplots reflecting topographical differences in within-network nodal dispersion between patients with TRD (red) and HCs (blue). **(C)** Violinplots reflecting topological differences in within-network nodal degree between patients with TRD and HCs. **(D)** Scatterplots reflecting the association between within-network nodal degree and within-network nodal dispersion separately for patients with TRD and HCs. Pearson’s correlation coefficients are reported below the scatterplots for each group separately, together with associated Fisher r-to-z tests for independent samples comparing the strength of the correlations across groups. CoN = Control Network; DAN = Dorsal Attention Network; DMN = Default Mode Network; HC = healthy controls; LiN = Limbic Network; SaN = Salience Network; SMN = Sensorimotor network; TRD = patients with treatment resistant depression; ViN = Visual Network. *p<0.05 FDR corrected, +p<0.05 uncorrected

### Within-network nodal degree

Comprehensively, the previous findings suggested that in TRD, nodes belonging to the same network are more integrated with each other. To confirm this hypothesis, we derived a measure of within-network modularity based on graph theoretical approaches. Within-network degree was derived for each intrinsic brain network, reflecting the ratio between existing edges a specific node shares with other nodes belonging to the same network and the total number of edges available in that network. Within-network nodal degree did not substantially differ between HC and TRD participants, except nodal degree decreased in the DMN and Sensorimotor Networks of patients with TRD (Figure 3C; p<0.05, uncorrected for multiple comparisons). However, when relating within-network nodal dispersion to within-network nodal degree, we consistently found a significant negative association between both nodal measures, particularly in TRD patients and to a lesser degree in HC (Figure 3D; p<0.05, FDR corrected for multiple comparisons if not reported otherwise, Pearson’s correlation coefficients and associated Fisher r-to-z tests for independent samples comparing the strength of correlations across groups reported in the plots). Notably, these findings were robust across distinct thresholds applied to generate the weighted connectivity matrices used to estimate nodal degree (Figure S4). In summary, these findings support the notion that decreased within-nodal dispersion, at least in patients, reflects increased within-network functional modularity. This negative association between nodal measures was prominent in TRD but not as developed in HCs, suggesting a more complex relationship between brain topology and cortical topography in the healthy human brain.

### DMN nodal dispersion and symptoms of depression

Finally, we aimed to associate typical symptoms of TRD with altered levels of within-network nodal dispersion and nodal degree. We focused only on changes affecting the DMN, given the recurrent association of this system with typical symptoms of major depression such as increased anxiety, depressed mood, and reduced mindfulness (11,23,24). In line with previous work, our patient sample showed increased levels of trait anxiety as measured through the STAI questionnaire (Figure 4A; p<0.05, FDR corrected for multiple comparisons). Trait anxiety correlated negatively with within-DMN nodal dispersion (R_TRD_ = -0.27; R_HC_ = 0.33; Fisher r-to-z tests for independent samples, z = -2.48) in TRD patients, but not in HCs, and positively with within-DMN nodal degree (R_TRD_ = 0.38; R_HC_ = 0.30; z = -0.36) in both groups (Figure 4B-C; p<0.05, FDR corrected for multiple comparisons). Similarly, depressive symptoms measured using the RSQ22 questionnaire were increased in TRD patients (Figure 4D; p<0.05, FDR corrected for multiple comparisons). Depressive symptoms showed a significant negative association with within-DMN nodal dispersion (R_TRD_ = -0.31; R_HC_ = 0.10; z = 1.73) in TRD patients, but not in HCs, while being positively linked to within-DMN nodal degree (R_TRD_ = 0.24; R_HC_ = 0.20; z = -0.17) in both groups (Figure 4E-F; p<0.05, FDR corrected for multiple comparisons if not specified otherwise). Mindfulness levels are often reduced in patients suffering from major depressive episodes (23,24) in line with the lower FFMQ scores found in our patient sample (Figure 4G; p<0.05, FDR corrected for multiple comparisons). Mindfulness scores were positively associated with within-DMN nodal dispersion (R_TRD_ = 0.31; R_HC_ = -0.37, z = -2.83) and negatively correlated with within-DMN nodal degree (R_TRD_ = -0.39; R_HC_ = -0.05; z = 1.45) in TRD patients but not in HCs (Figure 4E-F; p<0.05, FDR corrected for multiple comparisons). These findings suggest that, at least in TRD patients, reduced nodal dispersion and increased nodal degree among DMN regions are of a dysfunctional nature and may underlie common clinical symptoms of TRD.

**Figure 4.**
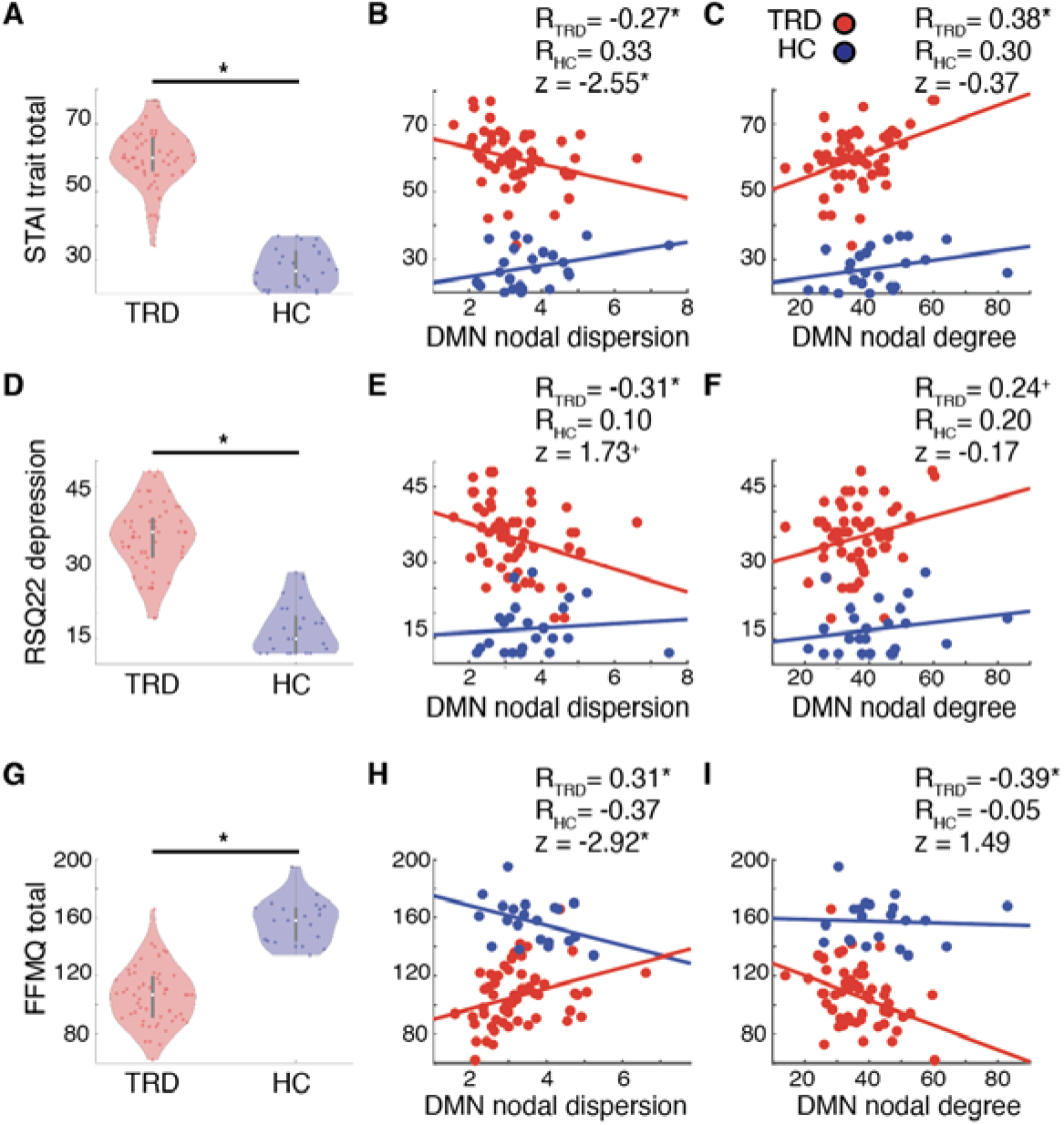
DMN nodal dispersion correlates with symptoms of depression. **(A** Levels of trait anxiety as measured through STAI trait total scores are significantly higher in patients with TRD (red violinplots) when compared to HCs (blue violinplots). A significant negative correlation is found between trait anxiety and within-DMN nodal dispersion in TRD but not in HC **(B)**, while trait anxiety shows a positive correlation to within-DMN nodal degree in both groups **(C)**. **(D)** Depressive symptoms as measured through RSQ22 total scores are significantly higher in patients with TRD (red violinplots) when compared to HCs (blue violinplots). A significant negative correlation is found between depressive symptoms and within-DMN nodal dispersion in TRD but not in HC **(E)**. Depressive symptoms show a positive correlation to within-DMN nodal degree in both groups **(F)**. **(G)** Levels of mindfulness as measured through FFMQ total scores are significantly lower in patients with TRD when compared to HCs. Mindfulness correlates positively with within-DMN nodal dispersion in TRD but not in HC **(H)**, while it correlates negatively with within-DMN nodal degree in TRD but not in HC **(I)**. DMN = Default Mode Network; HC = healthy controls; TRD = patients with treatment resistant depression. *p<0.05 FDR corrected, ^+^p<0.05 uncorrected

We finally assessed whether within-DMN nodal dispersion at baseline could predict clinical changes in TRD patients following an 8-week course of group therapy involving either the MBCT or HEP intervention. In this sample, both treatment arms did not significantly differ from each other regarding clinical changes over time, despite a previous study by our group using a larger sample that included the current one revealing a significant clinical advantage for MBCT relative to HEP (23,24). However, in line with the previous study, we found a main effect of time across both groups with improved STAI trait, FFMQ, and RSQ22 scores after 8 and 24 weeks (Figure S5 and Table S3). Contrary to our expectations, DMN nodal dispersion at baseline did not significantly predict clinical change scores at 8 or at 24 weeks (Table S4).

## Discussion

The brain is functionally organized along connectivity gradients that separate primary sensory and motor areas from transmodal associative cortices overlapping with the DMN. This study explored how TRD impacts this fundamental topography of hierarchical cortical organization. We capitalized on rs-fMRI data acquired in TRD patients and HCs and applied novel gradient extraction tools to assess gradient imbalances within major intrinsic brain networks. Although the global hierarchical architecture was similar across the two groups, we found that the brain regions belonging to the same network are located more closely to each other in topographical gradient space in TRD patients relative to HCs. Reduced within-network nodal dispersion correlated with higher levels of nodal degree derived through graph theory-based topology measures, overall suggesting higher within-network functional integration in TRD. Decreased DMN dispersoin in TRD patients correlated with higher current depression and anxiety, as well as reduced mindfulness. Overall, these findings suggest deleterious cortical network topography and topology in TRD and highlight an important role for the DMN in mediating core symptoms of depression.

### Increased within-network integration in TRD

The pervasive correlation between nodal degree and nodal dispersion in our patient sample suggests that TRD impacts cortical hierarchies by driving hyper-modularity within several brain networks (46). Other neuropsychiatric conditions have been shown to impact cortical connectivity gradients. Autism spectrum disorder has been shown to alter brain topography by showing atypical connectivity transitions between sensory and higher-order DMN regions (17). Our findings align with previous reports of altered cortical gradient organization in individuals with cognitive vulnerability to depression (19) and in a larger sample of patients with major depression (18). Individuals with cognitive vulnerability to depression have been shown to display reduced gradient scores in the left insula, which correlated with attentional scores commonly deficient in patients, suggesting that gradient reorganization may precede the onset of depression (19). A recent study involving a large sample of patients showed that major depressive disorder exhibits abnormal global topography of the principal primary-to-transmodal gradient (18). These focal alterations of gradient scores mostly affected transmodal areas implicated in higher-order cognition overlapping with the DMN (18).

### DMN modularity mediates symptoms of depression

Despite numerous efforts to map DMN dysfunctions in depression, important inconsistencies exist regarding the location and directionality of connectivity changes, with both hyper- (10), and hypoconnectivity findings reported in the literature (47). Several factors could contribute to these inconsistent findings, including differences in analytical approaches and heterogeneity among the recruited patients. Disease duration, perseverance of symptoms, and heterogenous subtypes of depression (8,9) may account for important sources of variability, as do head movement in the scanner, and differing data acquisition protocols and preprocessing pipelines (35–37). Overall, our findings align well with previous reports of DMN hyperconnectivity found in patients with depression (9,10). Hyperconnectivity among DMN regions in depression is consistent with our interpretation of reduced nodal dispersion reflecting increased within-network modularity. Several prior studies in both HCs and patients with depression have associated DMN hypersynchrony with self-referential processes affected in depression, including reduced mindfulness and social-emotional dysfunction (10,11,48), supporting the deleterious nature of DMN hyper-modualrity in TRD.

### Limitations and future directions

Three limitations need to be considered when interpreting our findings as potential evidence of within-network hyper-modularity in TRD. First, methods used to extract connectivity gradients may overly focus on cortical aspects of hierarchical brain organization, ignoring the influence of subcortical areas. Further efforts are needed to better assess the contribution of subcortical regions to predominant brain gradients and their dysfunction in neuropsychiatric diseases. Second, although findings of reduced within-network nodal dispersion were consistently found when using global signal regression or medium to high parcellated atlases, the method chosen to derive cortical connectivity gradients greatly influenced the outcome of the analyses. Third, and contrary to our expectations, DMN nodal dispersion in TRD did not predict improvements in clinical scores following either a mindfulness-based intervention or a health enhancement program. Given the recent discovery of distinct biotypes in major depressive disorder (8,9), longitudinal intervention studies involving larger patient samples with differing levels of clinical severity are needed to validate our findings of dysfunctional gradient architecture in TRD.

## Data Availability

Participants data is not publicly shared due to privacy concerns but is available from the corresponding author after reasonable request.

https://github.com/lollopasquini

## Acknowledgements

This work was supported by NIH grant K99-AG065457 to LP, DP2-MH119735 to MS, NIH/NCCAM grant R01-AT004572-02S1 to SJE and DHM. We thank the participants and their families for their invaluable contributions to depression research.

## Supplementary Information

### Supplementary Methods

#### Nodal dispersion

We derived a measure of within-network nodal dispersion for each participant by plotting the first three connectivity gradients against each other to derive a topographical three-dimensional Euclidean space. Within this manifold we then calculated the Euclidean distance between nodes belonging to the same intrinsic brain network (42) (Figure 1Cd). More formally, nodal dispersion was defined for each node as:

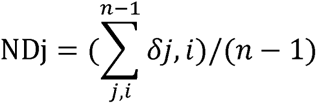

where NDj refers to the nodal dispersion of an individual node within a specific intrinsic brain network, e.g., the DMN, and n refers to the total number of nodes within this network. δj,i reflects de Euclidean distance between node j and another node i belonging to that network. δj,i is iteratively generated between a node and all other nodes in a network and eventually averaged. Nodal dispersion measures were derived for each node belonging to a specific intrinsic brain network and averaged across nodes, yielding a final estimate of within-network nodal dispersion for each participant. We performed several control analyses to assess the impact of methodological parameters on our findings. We derived measures of within-network nodal dispersion by using the first six gradients instead of three; global signal regression on rs-fMRI data; the Schaefer Altas with 1000 or 200 parcels; Laplacian embedding instead of diffusion embedding to derive cortical gradients; or angular normalization instead of cosine similarity to derive the dissimilarity matrices. Findings from these analyses are presented in the Supplementary Figures and Tables. Finally, we also derived a measure of between-network nodal dispersion calculated as the Euclidean distance between network centroids (i.e., the arithmetic mean of all nodes belonging to the same network).

#### Statistical analyses

Chi-square tests were used to compare sex, handedness, ethnicity, and race distributions across groups (p<0.05 uncorrected). Two sample t-tests were used to compare continuous demographical and clinical variables, functional connectivity matrices, and parcel-level gradient maps between HCs and patients (p<0.05 uncorrected). ANOVA models and associated post-hoc t-test were used to compare measures of within-network nodal dispersion and nodal degree across HCs and patients with TRD (p<0.05 FDR corrected for multiple comparisons if not specified otherwise). Multiple linear regression analyses were used to associate measures of within-network nodal dispersion to demographical variables and mean framewise head displacement (p<0.05 uncorrected). Pearson’s correlation analyses were used to associate measures of within-network nodal degree to measures of within-network nodal dispersion separately for each group (p<0.05 FDR corrected for multiple comparisons if not specified otherwise). To compare the strength of the correlations across groups, Fisher r-to-z transformation tests for independent samples were performed using an online calculator (https://www.psychometrica.de/correlation.html). Pearson’s correlation analyses were used to associate measures of within-DMN nodal degree and dispersion to neuropsychological questionnaire scores at baseline separately for each group (p<0.05 FDR corrected for multiple comparisons if not specified otherwise). Again, Fisher r-to-z tests for independent samples were performed to compare the strength of the correlations across groups. Three repeated measures ANOVA were performed to evaluate the effect of MBCT and HEP interventions over time on STAI trait, RSQ22, and FFMQ scores of patients with TRD. Pearson’s correlation analyses were used to associate within-DMN nodal dispersion to change in STAI trait, RSQ22, and FFMQ scores at 8 and 24 weeks after completing the interventions. Both intervention arms were combined into one sample for this last set of analyses assessing the association between longitudinal change scores and DMN functional measures at baseline.

### Supplementary Figures and Tables

**Supplementary Figure S1.**
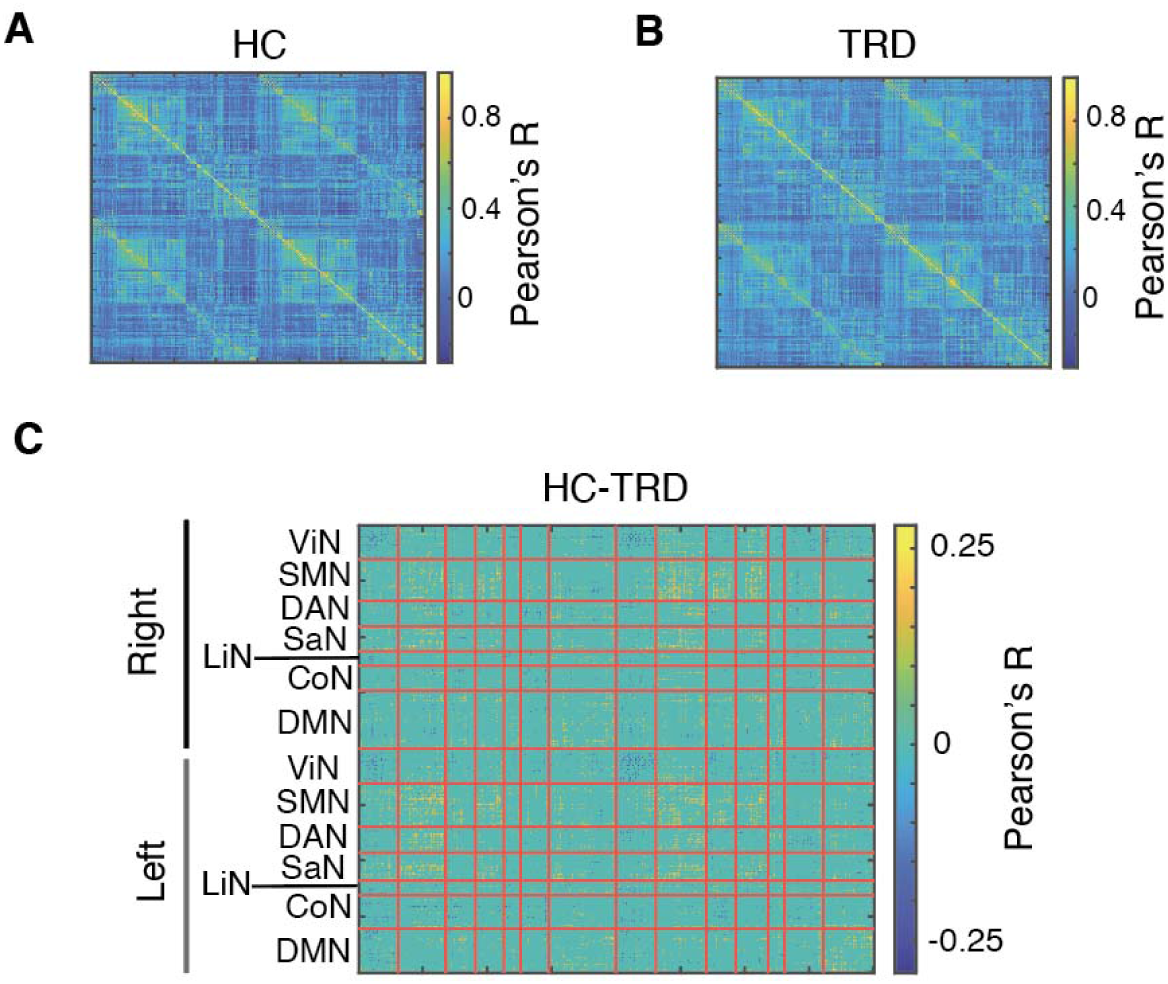
Functional connectivity matrices derived from rs-fMRI data of **(A)** HCs and **(B)** patients with TRD. **(C)** Subtraction matrix showing functional connectivity differences across both groups. Warm colors reflect connectivity decreases in patients, while cold colors reflect connectivity increases in patients (p<0.05 uncorrected, no group differences survived FDR multiple comparison correction). CoN = Control Network; DAN = Dorsal Attention Network; DMN = Default Mode Network; HC = healthy controls; LiN = Limbic Network; SaN = Salience Network; SMN = Sensorimotor network; TRD = patients with treatment resistant depression; ViN = Visual Network.

**Supplementary Figure S2.**
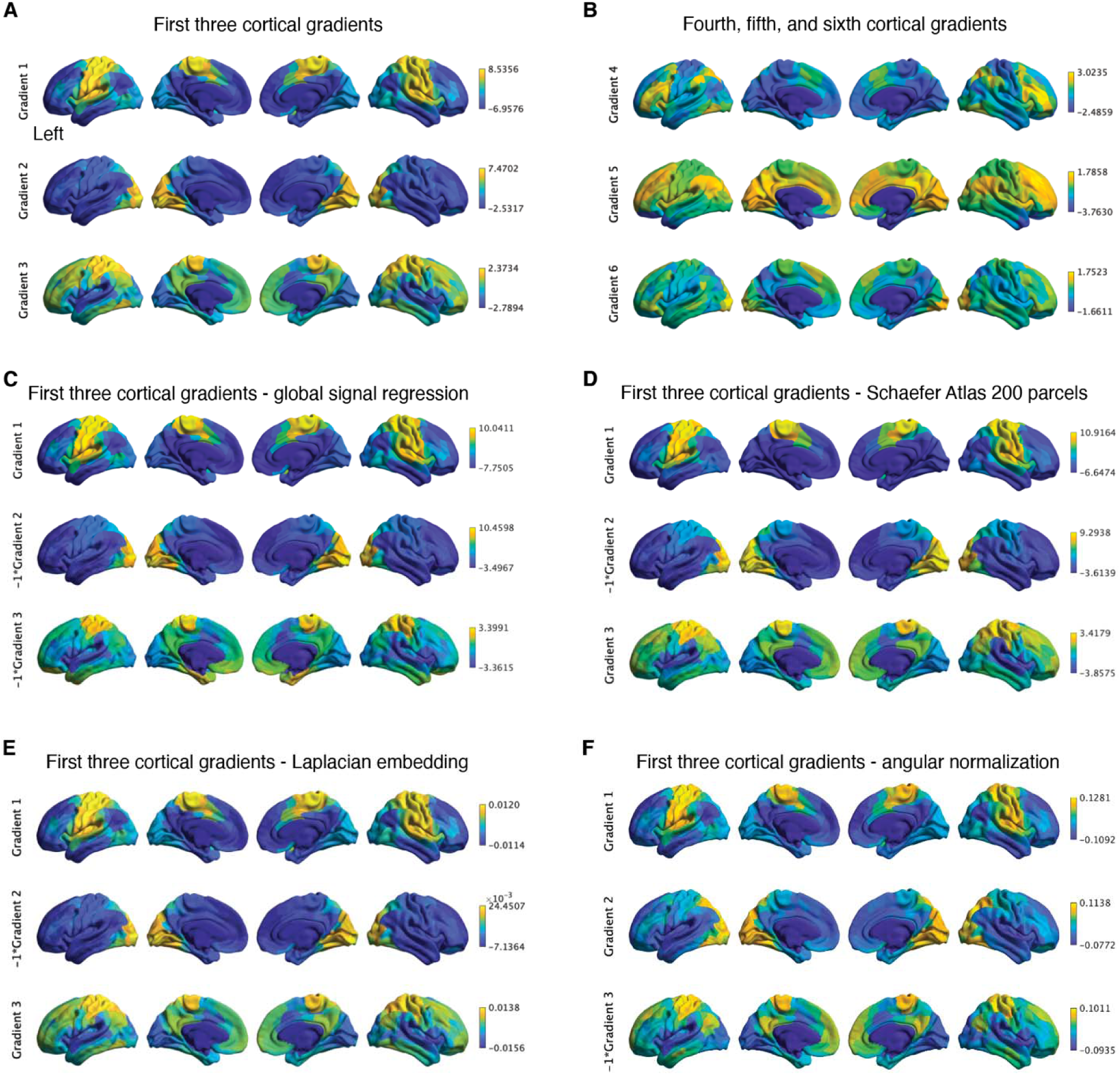
Cortical gradient extraction control analyses. **(A)** Maps of cortical Gradients 1-3 as shown in the main findings. These gradients explained most of the variance in connectivity, followed by Gradients 5-6 **(B)**. First three cortical Gradients derived either through **(C)** global signal regressed rs-fMRI data; **(D)** Schaefer atlas with 200 parcels; **(E)** Laplacian embedding; or **(F)** angular normalization to generate the dissimilarity matrices. The sing assigned to gradients by the decomposition algorithm is arbitrary; we flipped the sign of some Gradients to better visualize the spatial similarity across corresponding gradient maps.

**Supplementary Figure S3.**
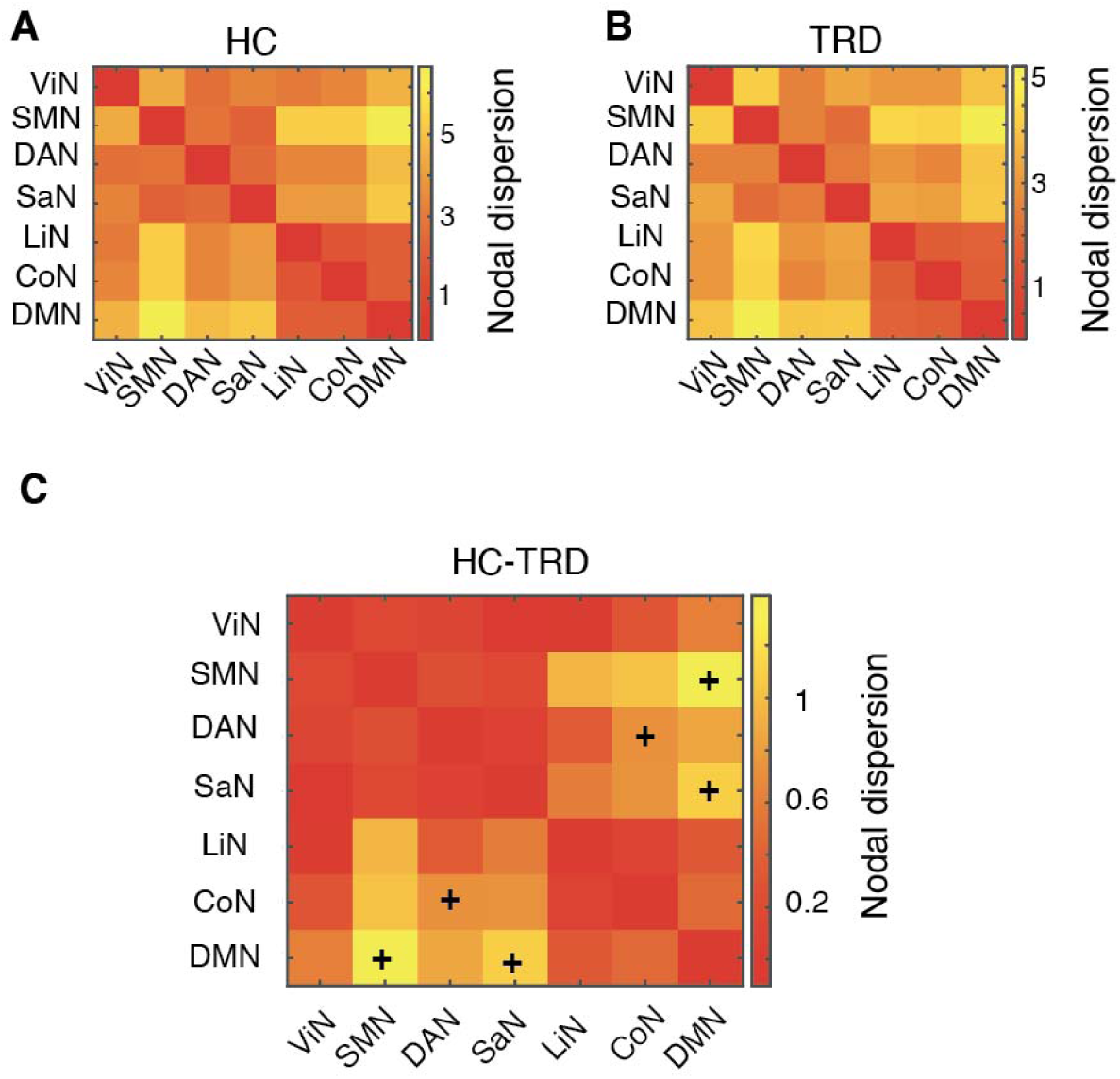
Between-network nodal dispersion. Between-network nodal distance in **(A)** HCs and **(B)** patients with TRD. **(C)** Significant reductions in between-network nodal dispersion were found in patients with TRD, affecting the Sensorimotor and DMN, the Salience and DMN, and the Control and Dorsal Attention Network. None of these findings survived FDR correction for multiple comparisons. +p<0.05 uncorrected. CoN = Control Network; DAN = Dorsal Attention Network; DMN = Default Mode Network; HC = healthy controls; LiN = Limbic Network; SaN = Salience Network; SMN = Sensorimotor network; TRD = patients with treatment resistant depression; ViN = Visual Network.

**Supplementary Figure S4.**
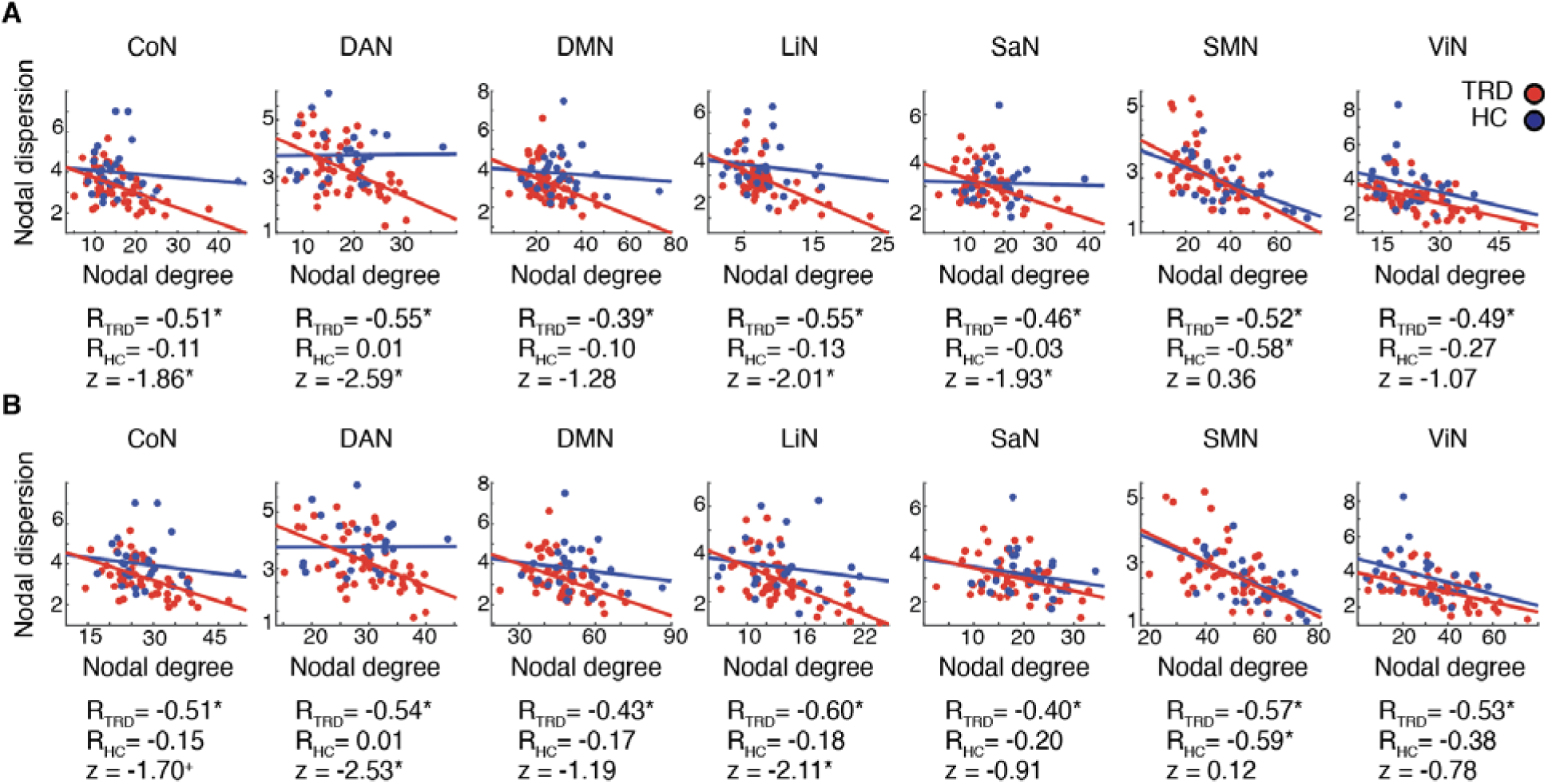
Within-network nodal dispersion and nodal degree control analyses. Scatterplots reflecting the association between within-network nodal degree and within-network nodal dispersion separately for patients with TRD and HCs. Within-network nodal degree was repeatedly estimated by applying distinct thresholds when generating the weighted connectivity matrices. **(A)** Connectivity threshold set at 0.45. **(B)** Connectivity threshold set at 0.25. Pearson’s correlation coefficients are reported below the scatterplots for each group separately together with associated Fisher r-to-z tests for independent samples comparing the strength of the correlations across groups. CoN = Control Network; DAN = Dorsal Attention Network; DMN = Default Mode Network; HC = healthy controls; LiN = Limbic Network; SaN = Salience Network; SMN = Sensorimotor network; TRD = patients with treatment resistant depression; ViN = Visual Network. *p<0.05 FDR corrected, ^+^p<0.05 uncorrected

**Supplementary Figure S5.**
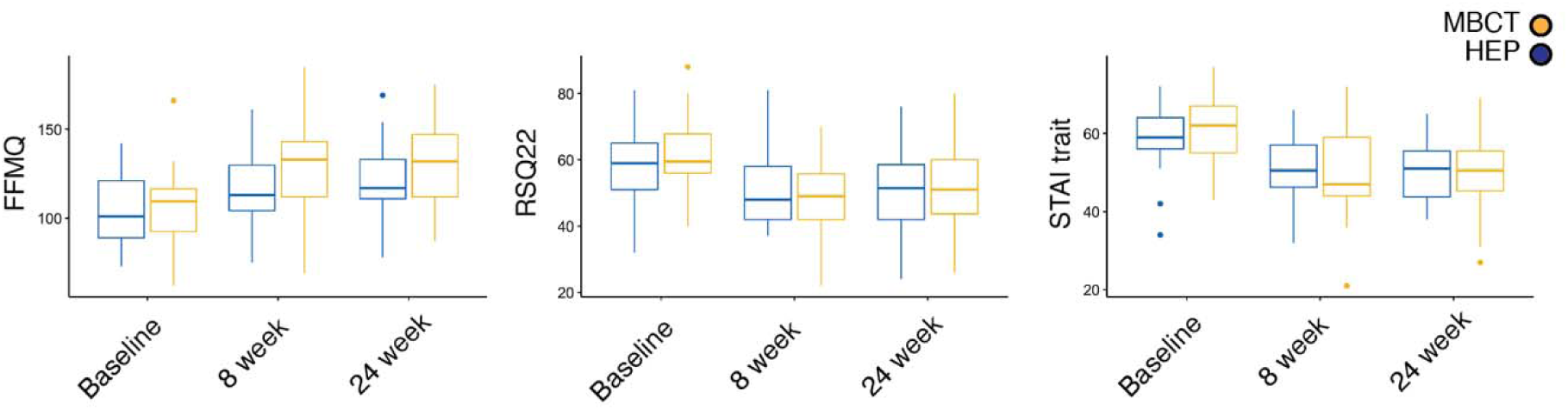
Changes in clinical scores with MBCT and HEP interventions. Patients with TRD report higher levels of mindfulness, lower levels of depression, and lower levels of trait anxiety at 8 and 24 weeks after completing a MBCT (orange) or a HEP intervention (blue). HEP = health enhancement program; FFMQ = Five Facet; MBCT = Mindfulness Questionnaire; Mindfulness-Based Cognitive Therapy; RSQ22 = Nolen-Hoeksema’s Response Styles Questionnaire; STAI = State-Trait Anxiety Inventory.

**Supplementary Table S1.** Within-network nodal dispersion is not associated with head movement, age, or sex. Seven separate multiple linear regression models were run in patients with TRD to assess the link between within-network nodal dispersion, used as the dependent variable, and mean frame-wise head displacement (FD), age, and sex. There were no significant associations. CoN = Control Network; DAN = Dorsal Attention Network; DMN = Default Mode Network; LiN = Limbic Network; SaN = Salience Network; SMN = Sensorimotor network; TRD = patients with treatment resistant depression; ViN = Visual Network.

|  | Within-network nodal dispersion |  |  |  |  |  |  |
| --- | --- | --- | --- | --- | --- | --- | --- |
|  | CoN | DAN | DMN | LiN | SaN | SMN | ViN |
| **Age** | $\beta = -0.02$<br>$p = 0.17$ | $\beta = 0.00$<br>$p = 0.95$ | $\beta = -0.01$<br>$p = 0.51$ | $\beta = 0.00$<br>$p = 0.76$ | $\beta = 0.00$<br>$p = 0.86$ | $\beta = -0.01$<br>$p = 0.56$ | $\beta = 0.00$<br>$p = 0.83$ |
| **Sex** | $\beta = 0.16$<br>$p = 0.58$ | $\beta = 0.06$<br>$p = 0.86$ | $\beta = 0.19$<br>$p = 0.54$ | $\beta = -0.25$<br>$p = 0.43$ | $\beta = -0.18$<br>$p = 0.48$ | $\beta = -0.04$<br>$p = 0.90$ | $\beta = 0.17$<br>$p = 0.55$ |
| **FD** | $\beta = 0.03$<br>$p = 0.98$ | $\beta = -0.83$<br>$p = 0.49$ | $\beta = 0.92$<br>$p = 0.44$ | $\beta = -1.04$<br>$p = 0.40$ | $\beta = -0.44$<br>$p = 0.66$ | $\beta = 0.39$<br>$p = 0.73$ | $\beta = -1.58$<br>$p = 0.14$ |

**Supplementary Table S2.** Within-network nodal dispersion across methodological parameters. Group-mean within-network nodal dispersion and standard deviation in brackets. We systematically assessed whether group differences in within-network nodal dispersion were affected by methodological parameters, such as: (i) including up to six cortical gradients when assessing the Euclidean distance between nodes; (ii) performing global signal regression on rs-fMRI data; (iii) using the Schaefer Atlas at a lower spatial resolution of 200 parcels; (iv) using the Schaefer Atlas at a higher spatial resolution of 1000 parcels; (v) applying Laplacian embedding to derive cortical gradients; or (vi) using angular normalization to generate the dissimilarity matrices. Raw p values are reported; *denotes p<0.05 FDR corrected for multiple comparisons. Significant FDR corrected reductions in within-network nodal degree in patients are highlighted in the orange cells. CoN = Control Network; DAN = Dorsal Attention Network; DMN = Default Mode Network; LiN = Limbic Network; SaN = Salience Network; SMN = Sensorimotor network; TRD = patients with treatment resistant depression; ViN = Visual Network.

| i. | First six cortical gradients |  |  |  |  |  |  |
| --- | --- | --- | --- | --- | --- | --- | --- |
|  | ViN | SMN | DAN | SaN | LiN | CoN | DMN |
| TRD | 3.7 (0.9) | 3.2 (0.9) | 3.9 (0.8) | 3.7 (0.8) | 3.6 (1.0) | 4.1 (0.8) | 4.2 (1.0) |
| HC | 4.5 (1.3) | 2.8 (0.9) | 4.3 (0.8) | 3.6 (0.9) | 4.0 (1.1) | 4.7 (1.1) | 4.5 (1.1) |
| p | 0.003* | 0.074 | 0.025 | 0.850 | 0.083 | 0.010* | 0.155 |
| ii. | Global signal regression |  |  |  |  |  |  |
|  | ViN | SMN | DAN | SaN | LiN | CoN | DMN |
| TRD | 3.5 (0.9) | 2.8 (0.8) | 3.6 (0.9) | 3.1 (0.7) | 3.3 (1.1) | 3.8 (1.0) | 3.7 (1.0) |
| HC | 4.3 (1.6) | 2.6 (0.8) | 4.3 (1.0) | 3.5 (0.9) | 4.2 (1.5) | 4.6 (1.7) | 4.3 (1.3) |
| p | 0.007* | 0.601 | 0.002* | 0.026* | 0.003* | 0.005* | 0.013* |
| iii. | Schaefer Atlas 200 parcels |  |  |  |  |  |  |
|  | ViN | SMN | DAN | SaN | LiN | CoN | DMN |
| TRD | 3.6 (1.3) | 3.2 (1.4) | 4.0 (1.5) | 3.8 (1.2) | 3.4 (1.5) | 4.3 (1.4) | 4.0 (1.4) |
| HC | 4.5 (1.7) | 2.9 (1.2) | 4.4 (1.3) | 3.5 (1.4) | 3.7 (1.5) | 4.7 (1.6) | 4.2 (1.2) |
| p | 0.007* | 0.299 | 0.227 | 0.401 | 0.353 | 0.283 | 0.416 |
| iv. | Schaefer Atlas 1000 parcels |  |  |  |  |  |  |
|  | ViN | SMN | DAN | SaN | LiN | CoN | DMN |
| TRD | 2.8 (0.9) | 2.1 (0.6) | 2.9 (0.7) | 2.5 (0.6) | 2.5 (0.8) | 3.0 (0.8) | 2.9 (0.8) |
| HC | 3.4 (1.1) | 2.0 (0.6) | 3.4 (0.8) | 2.7 (0.7) | 3.1 (1.0) | 3.6 (1.1) | 3.2 (0.8) |
| p | 0.002* | 0.562 | 0.003* | 0.218 | 0.004* | 0.013* | 0.043 |
| v. | Laplacian embedding – values multiplied by 100 |  |  |  |  |  |  |
|  | ViN | SMN | DAN | SaN | LiN | CoN | DMN |
| TRD | 1.3 (0.2) | 1.3 (0.2) | 1.3 (0.2) | 1.3 (0.3) | 1.2 (0.2) | 1.3 (0.2) | 1.2 (0.2) |
| HC | 1.4 (0.2) | 1.3 (0.3) | 1.3 (0.2) | 1.3 (0.2) | 1.2 (0.2) | 1.3 (0.2) | 1.2 (0.2) |
| p | 0.080 | 0.150 | 0.266 | 0.379 | 0.514 | 0.968 | 0.234 |
| vi. | Angular normalization – values multiplied by 100 |  |  |  |  |  |  |
|  | ViN | SMN | DAN | SaN | LiN | CoN | DMN |
| TRD | 6.8 (1.0) | 7.3 (1.2) | 7.9 (1.0) | 7.2 (1.0) | 5.9 (1.3) | 7.3 (0.9) | 7.1 (1.0) |
| HC | 6.9 (1.1) | 7.5 (1.3) | 8.4 (1.1) | 7.5 (1.1) | 6.0 (1.1) | 7.5 (1.2) | 7.5 (1.1) |
| **P** | 0.657 | 0.440 | 0.051 | 0.214 | 0.601 | 0.337 | 0.069 |

**Supplementary Table S3.** Repeated measurement ANOVA models assessing change in STAI trait, RSQ22, and FFMQ scores in patients. Repeated measurement ANOVA models revealed a significant effect of time on scores of STAI trait, RSQ22, and FFMQ from baseline to 8 and 24 weeks. No significant group-time interactions were found. FFMQ = Five Facet Mindfulness Questionnaire; RSQ22 = Nolen-Hoeksema’s Response Styles Questionnaire; STAI = State-Trait Anxiety Inventory.

| **Change score** | **Group** | **Time** | **Group\*Time** |
| --- | --- | --- | --- |
| **STAI trait** | F = 0.0<br>p = 0.96 | F = 19.2<br>p < 0.001 | F = 0.5<br>p = 0.61 |
| **RSQ22** | F = 0.10<br>p = 0.78 | F = 10.9<br>p < 0.001 | F = 0.6<br>p = 0.57 |
| **FFMQ** | F = 4.0<br>p < 0.05 | F = 11.6<br>p < 0.001 | F = 1.0<br>p = 0.36 |

**Supplementary Table S4.** Within-DMN nodal dispersion and change in STAI trait, RSQ22, and FFMQ scores in patients. Within-DMN nodal dispersion at baseline was correlated with change in STAI trait, RSQ22, and FFMQ scores at 8 and 24 weeks after the intervention. Since our analyzes revealed a main effect of time on clinical scores but not a time x group interaction, these correlation analyses were performed by combining in one group patients undergoing Mindfulness-Based Cognitive Therapy and the health enhancement program. There were no significant associations. FFMQ = Five Facet Mindfulness Questionnaire; RSQ22 = Nolen-Hoeksema’s Response Styles Questionnaire; STAI = State-Trait Anxiety Inventory.

| Within-DMN nodal dispersion at baseline |  |  |
| --- | --- | --- |
| Change score | 8 weeks - baseline | 24 weeks - baseline |
| **STAI trait** | R = 0.11<br>p = 0.42 | R = 0.01<br>p = 0.92 |
| **RSQ22** | R = 0.05<br>p = 0.74 | R = 0.00<br>p = 0.95 |
| **FFMQ** | R = 0.01<br>p = 0.95 | R = 0.10<br>p = 0.49 |

